# Genome-wide study of somatic symptom and related disorders identifies novel genomic loci and map genetic architecture

**DOI:** 10.1101/2025.07.16.25331639

**Authors:** Vera Fominykh, Piotr Jaholkowski, Alexey A. Shadrin, Elise Koch, Laura B. Luitva, Dorte H. Mikkelsen, Mischa Lundberg, Ole Birger Pedersen, Sisse Rye Ostrowski, Christian Erikstrup, Maria Didriksen, Christina Mikkelsen, Erik Sørensen, Henrik Ullum, Mie Topholm Bruun, Bitten Aagaard, Kaarina Kowalec, Robert Karlsson, Håkan Karlsson, Christina Dalman, Pravesh Parekh, Viktoria Birkenæs, Yury Seliverstov, Bernhard Landwehrmeyer, Ida E. Sønderby, DBDS genetic consortium, Estonian Biobank research team, Olav B. Smeland, Kevin S. O’Connell, Lu Yi, Patrick F. Sullivan, Thomas M. Werge, Lili Milani, Ole A. Andreassen

## Abstract

Somatic symptom and related disorders (SSRD) are characterized by a mixture of neurological and psychiatric features and include functional neurological (FND) and somatic symptom disorders (SomD). While these complex neuropsychiatric disorders show evidence of genetic susceptibility, there are no genome-wide association studies (GWAS) of SSRD, and the heritability is unknown.

We did a GWAS of a total of 22,203 patients with SSRD, and 1,831,107 controls of European ancestry. We identified one genome-wide significant locus (chromosome 8:65565084) in SSRD, and one additional locus (chromosome 16:49074278) in the SomD subgroup (n cases = 18,536). The observed-scale SNP heritability was estimated to be 7.3 % for SSRD, 15.7 % for FND and 7.7 % for SomD. FND and SomD were strongly genetically correlated (rg=0.94, SE=0.11, p=3.9E-18). SSRD showed significant genetic correlation with psychiatric disorders (highest with anxiety, post-traumatic stress disorders, depression, rg=0.3- 0.8), neurological disorders (migraine, chronic pain, rg=0.4-0.6) and immune-related diseases (rg=0.2-0.3). Functional follow-up analysis of SSRD loci implicated the genes *CYP7B1, BHLHE22,* and *CBLN1,* which are involved in metabolic and brain-related processes, suggesting common underlying pathways.

We identified genomic loci associations with SSRD and showed strong genetic correlation between FND and SomD and with neurological and psychiatric disorders, as well as immune-related diseases. The current findings highlight shared underlying pathophysiological processes between SSRD diagnostic categories.

## Introduction

Somatic symptom and related disorders (SSRD), including functional neurological disorders (FND) and somatoform disorders (SomD), are highly prevalent [Finkelstein SA. et al., 2025] and characterised by somatic symptoms associated with significant distress or impairment. These disorders involve a complex interplay of psychological, social, and biological factors, and often co-occurring mental health conditions. Despite some recent advances in understanding their pathophysiology, the disease mechanisms remain unclear [Galli S. et al., 2020, Pellicciari R. et al., 2014; Spagnolo P. et al., 2020, 2023].

The SSRD encompass FND and SomD, which are sometimes placed between the neurological and psychiatric areas of expertise and have been previously defined as “non-organic pathology” [Stone J. et al, 2017; Bell W. et al., 2020]. The FND are characterized by a range of clinical symptoms that mimic neurological conditions such as tremor, dystonia, seizures, and cognitive dysfunction, which often present together in complex combinations. They develop due to dysfunction of certain brain networks in the absence of identifiable organic pathology [Hallett M. et al.; 2022, Stone J. et al, 2017; Edwards MJ. et al., 2023]. SomD or somatic symptom disorders (DSM-5) are characterized by physical symptoms that are distressing and affect daily life and cannot be explained by coexisting organic conditions. These symptoms can manifest in various ways, including pain, fatigue, and gastrointestinal problems, with an impact on the quality of life [Stone J. et al., 2020].

Although SSRDs are the second most common reason for outpatient neurology consultations [Carson A. et al., 2011], have a significant impact on healthcare and societal costs [Carson A. et al., 2016], and have been described as a “crisis for neurology” [Stone J. et al., 2010; Perjoc RS. et al., 2023], the pathobiological underpinnings of these disorders are still mainly unknown [Serranová T. et al., 2024]. Brain imaging and neurophysiology have provided insights into the mechanisms behind the symptoms [Stone J. et al., 2014; Levenson et al., 2024], highlighting dysfunction in multimodal neuronal integration of information, which partially overlaps with psychiatric disorders [Patron VG. et al., 2022; Burton C. et al., 2020]. The overlapping clinical characteristics of SomD and FND indicate shared pathophysiology, and they are hypothesized to be genetically related [Patron VG. et al, 2022; Helgeland H. et al., 2025; Burton C. et al., 2020; Kurki M. et al., 2023, https://risteys.finngen.fi/endpoints/]. This is in line with the overlapping genetic architecture of neurological and psychiatric disorders [Smeland OB. et al, 2025].

The role of genetics in SSRD remains unclear. Some studies suggest that a positive family history correlates with higher morbidity risk among relatives of those with FND [Stamelou M. et al., 2013; Ljungberg L., 1957]. The existing genetic studies of SSRD [Asadi-Pooya AA. et al, 2016] are based on small sample sizes (n<100) and candidate gene approaches [Spagnola P. et al., 2020, 2023; Weber S. et al., 2024; Leu C. et al., 2020; Apazoglou K. et al., 2018; Jungilligens J. et al., 2022]. In fact, the number of studies on SSRD genetics is realtively low compared to other disorders (see Supplementary (Suppl.) Fig. 1). No adequately powered genome-wide association studies (GWAS) have been conducted in SSRD or subtypes FND and SomD: the SNP heritability remains unknown, and the underlying molecular-genetic pathways have not been identified yet.

**Fig. 1.**
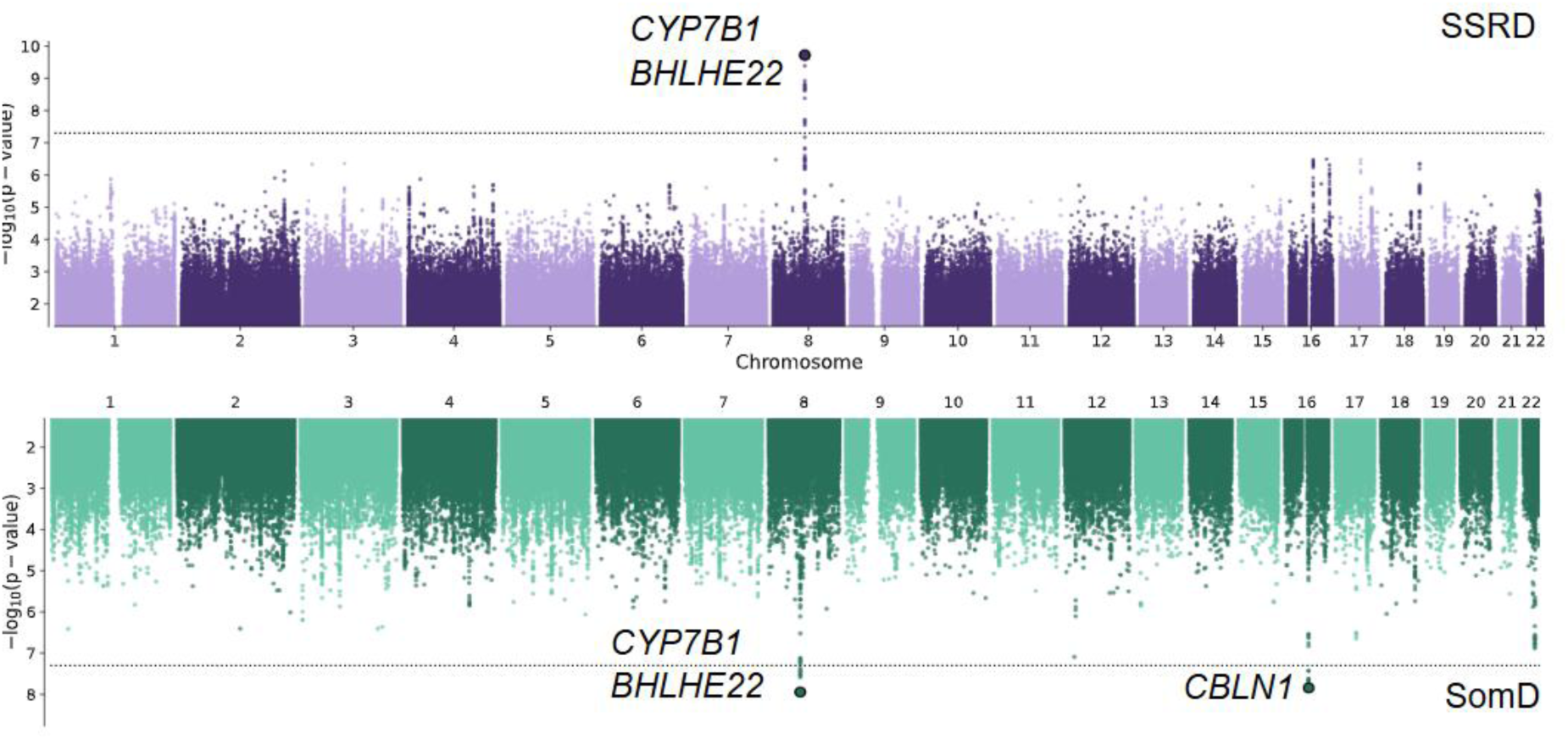
Manhattan plot for the genome-wide association analysis (GWAS) of the combined meta-nalysis (purple plot) on somatic-symptoms related disorders (SSRD, combined functional neurological disorders (FND) and somatoform disorders (SomD)), and the separate GWAS on SomD (green plot). −log10 *p*-values are plotted for all variants across chromosomes 1–22. The bold dots indicate loci with a lead variant genome-wide association study *p*-value of < 5×10^−8^. The grey line indicates the threshold for genome-wide significance (*p*-value of 5×10^−8^).

In the current study, we performed a GWAS meta-analysis involving a total of 22,203 SSRD patients, along with 1,831,107 controls. We aimed to identify genomic loci associated with these phenotypes, explore the genetic correlation and heritability, investigate potential genetic links with psychiatric, neurological, and somatic conditions, and identify molecular genetic pathways with functional annotation methods.

## Materials and methods

### Cohorts

The GWAS discovery meta-analysis comprised n=22,203 SSRD patients and 1,831,107 controls across seven cohorts. The cases included n=4,269 FND patients and n=18,536 SomD patients. All participants were of European ancestry (Table 1). We meta-analysed GWAS data from five Nordic countries – Norway, Denmark, Estonia, Iceland, and Finland, along with the United Kingdom (UK), and the USA. The dataset included the Norwegian Mother, Father and Child Cohort Study (MoBa, Norway, Magnus P. et al., 2016), Copenhagen Hospital Biobank (CHB) and The Danish Blood Donor Study (DBDS) (Denmark, Erikstrup C. et al., 2023), Estonian Biobank (Estonia, Milani L. et al., 2024), deCODE (Iceland, Jónsson H. et al., 2017), FinnGen (Finland, Kurki M. et al., 2023), UK Biobank (UKB, United Kingdom, Sudlow C. et al., 2015; Bycroft C. et al, 2018), and All of Us (USA, Ramirez AH. et al., 2022; All of us, 2024). For the replication phase and validation of findings, we used summary statistics from an independent Swedish sample (Sweden Schizophrenia Study, Lichtenstein P. et al., 2006).

**Table 1.** Study summary for cases and controls.

| **Source** | **SSRD\* cases (n)** | **Controls (n)** | **FND cases (n)** | **Controls for FND (n)** | **SomD cases (n)** | **Controls for SomD (n)** |
| --- | --- | --- | --- | --- | --- | --- |
| **MoBa** | 1,614 | 202,810 | 308 | 204,721 | 1,353 | 203,203 |
| **UK Biobank** | 555 | 458,009 | 138 | 458,681 | 421 | 458,251 |
| **FinnGen** | 8,448 | 444,414 | 1,525 | 444,414 | 7,148 | 444,414 |
| **Estonian Biobank** | 4,960 | 172,672 | 506 | 205,244 | 4,530 | 173,222 |
| **Iceland, deCODE** | 2,931 | 341,030 | 785 | 325,403 | 2,265 | 341,147 |
| **Denmark** | 1,780 | 95,444 | 538 | 95,637 | 1,315 | 95,491 |
| **All of US (USA)** | 1,915 | 116,728 | 469 | 118,174 | 1,504 | 119,219 |
| **Sum:** | **22,203** | **1,831,107** | **4,269** | **1,852,274** | **18,536** | **1,834,947** |
| **N effective\*\*** |  | **87,220** |  | **17,021** |  | **72,986** |

\*SSRD (combined FND+SomD) number of cases corrected according to FND/SomD cases overlap. For FinnGen, we present the total number of FND+SomD cases with the subtracted shared cases according to publicly available information on the Risteys data browser. For details, see the Suppl. and Methods.
\*\*The effective sample size (N effective) was calculated using the formula $4 / (1/n \text{ cases} + 1/n \text{ controls})$ for each cohort. For the meta-analysis, the overall N effective was determined by summing the individual N effective values from all contributing cohorts.

SSRD cases were defined as having a lifetime main diagnosis according to ICD-10 criteria of FND (F44) or SomD (F45) (https://www.cdc.gov/nchs/icd/icd-10/, MOBA, UKB, Danish sample). If it was not possible to specify whether the patient had a primary or secondary diagnosis, we used the criteria that the diagnosis should have been made at a specialized healthcare facility (Estonia) or used available data for diagnosis of interest elsewhere (FinnGene, deCODE, All of Us). Participants from the same cohorts who did not have a primary or secondary diagnosis of FND or SomD were designated as control subjects. Detailed descriptions of the samples are provided in Table 1 and the Suppl. material. We also included information about comorbidity in Suppl. Table 1. For the SSRD sample GWAS analysis, cases were defined as having an FND AND/OR SomD diagnosis, accounting for overlapping comorbidity (except FinnGen, where this information was not available). The regional ethics committee of Norway approved the current study, and local ethics committees approved each of the studies included in the meta-analysis. All participants provided written informed consent.

### Genotyping, GWAS and meta-analysis

The genotyping, imputation and GWAS methodology details for each cohort are available in the Suppl. Inverse variance-based meta-analysis of the seven case-control GWAS cohorts was performed using the METAL software [Willer CJ. et al., 2010]. Genome-wide significance was set at a *p*-value threshold of 5×10^−8^. For the meta-analysis of SSRD phenotypes, we used the METAL tool with the overlap function “on” to reduce the effect of overlapping subjects [for details, see <u>METAL Documentation - Genome Analysis Wiki</u>], as the combined FinnGen FND+SomD GWAS with overlap accounting was not available. Before commencing meta-analysis, all GWAS summary statistics underwent uniform quality control and were harmonized using the v1.6.0 *cleansumstats* pipeline <u>GitHub_BioPsyk/cleansumstats</u> [Gadin JR. et al., 2023], and <u>precimed/python-convert</u> was used for creating Manhattan plots and QQ plots. Markers that exist only in one cohort were excluded from the meta-analysis. We used R (version 4.2.3) and git/bash Linux for simple data processing during data preparation and analysis. All results were provided on the GRCh37 build.

### Genetic correlation and heritability: LD score regression and SumHer/LD adjusted kinships (LDAK)

SNP-based heritability for SSRD, FND, and SomD was calculated using the LD Score regression method [LDSC, Bulik-Sullivan B. et al., 2015] on the observed scale. Liability scale heritability was not estimated, as the population prevalence for SSRD, FND, and SomD has an extensive range. LDSC genetic correlation analysis was performed to investigate a possible genetic overlap between FND and SomD, as well as the phenotypes of interest and other psychiatric, neurological, and immune-mediated disorders. GWAS summary statistics [Suppl. Table 2] were used to calculate genetic correlation between SSRD, FND, SomD, and major psychiatric disorders (bipolar disorder – BD [O’Connell KS. et al., 2025], major depressive disorder – MDD [Major Depressive Disorder Working Group of the Psychiatric Genomics Consortium et al., 2025], schizophrenia – SCZ [Trubetskoy V. et al., 2022]), the most often comorbid psychiatric disorders (anxiety, Purves KL. et al., 2020, attention deficit hyperactivity disorder (ADHD, Demontis D. et al., 2023), post-traumatic stress-related disorder (PTSD, Nievergelt CM. et al., 2024) and traits (neuroticism, Nagel M. et al., 2018), neurological disease (epilepsy, focal and generalised, ILAE, 2023, migraine [Bjornsdottir G. et al., 2023], multiple sclerosis [International Multiple Sclerosis Genetics Consortium, 2019]), chronic widespread pain [Zorina-Lichtenwalter K. et al., 2023], irritable bowel syndrome (IBS, Eijsbouts C. et al., 2021) and immune-related disorders (psoriasis, type 1 diabetes, systemic lupus erythematosus, systemic sclerosis, Sjogren’s syndrome, rheumatoid arthritis, autoimmune thyroid disorders, Crohn’s disease, and ulcerative colitis [for the source Suppl. Table 2, Fominykh V. et al., 2025]). The resulting *p*-values were Bonferroni corrected for multiple testing (alpha level 0.0011), considering the number of investigated phenotypes.

To support the LDSC estimates of the SNP-based heritability for FND and SomD, as well as the correlation between them, additional analysis was performed using the SumHer tool, as implemented in LDAK v6.1 [Speed D. et al., 2019], which allows the user to specify distinct heritability models. We used the Human Default Model with a power parameter set to -0.25, in which the contribution of each SNP to heritability depends on its MAF [Speed D. et al., 2019]. Genotypes of 12,926,669 common variants from 5,000 randomly selected unrelated European individuals from the UK Biobank were used as a reference panel to calculate SNP taggings. Variants that individually explain more than 1% of phenotypic variance were excluded from the analysis (LDAK option: --cutoff 0.01). We also did not apply genomic control (LDAK option: --genomic-control NO) as recommended by the LDAK authors [Speed D et al., 2019].

### Follow-up analysis (validation analysis)

For validation analysis, we used the Sweden Schizophrenia Study (Suppl.), SSRD n cases = 342, n controls = 11,340, FND: n cases = 87; n controls = 2,239. SomD: n cases = 128; n controls = 6,608. We examined the significance of the identified lead variants and effect direction in this independent Swedish cohort.

### Post-GWAS functional analysis

We applied FUMA v1.6.1 protocol for the identification of genomic loci and gene mapping strategies (positional mapping according to FUMA with 10 kb window size) and supported it by the Open Targets modelling for the annotation [Ghoussaini M et al., 2021, database version from October 2022, Watanabe 2017, de Leeuw CA et al., 2015]. We applied MAGMA v1.08 with default parameters as implemented in FUMA v1.6.1 [Watanabe 2017, de Leeuw CA et al., 2015] to perform gene-set analysis (17023 gene sets from MsigDB v2023.1Hs) and tissue specificity analysis (54 different tissue types from GTEx eQTL v8). Briefly, the meta-analysed summary statistics for SSRD, FND, and SomD GWAS data were used as input. The *p*-value threshold of 5×10^−8^ was used to define lead SNPs. An r^2^ of ≥0.6 and a genomic window of ± 250 kb were used to determine linkage disequilibrium (LD) with independent lead SNPs. *P*-values were corrected using the Bonferroni procedure following the standard protocol in FUMA.

## Results

The present study describes the largest up-to-date GWAS meta-analyses of 22,203 SSRD individuals and 1,831,107 controls (Table 1, and Suppl. Table 1 and Suppl. description, QQ plot for all GWASes are available in Suppl. Fig.2).

**Fig. 2.**
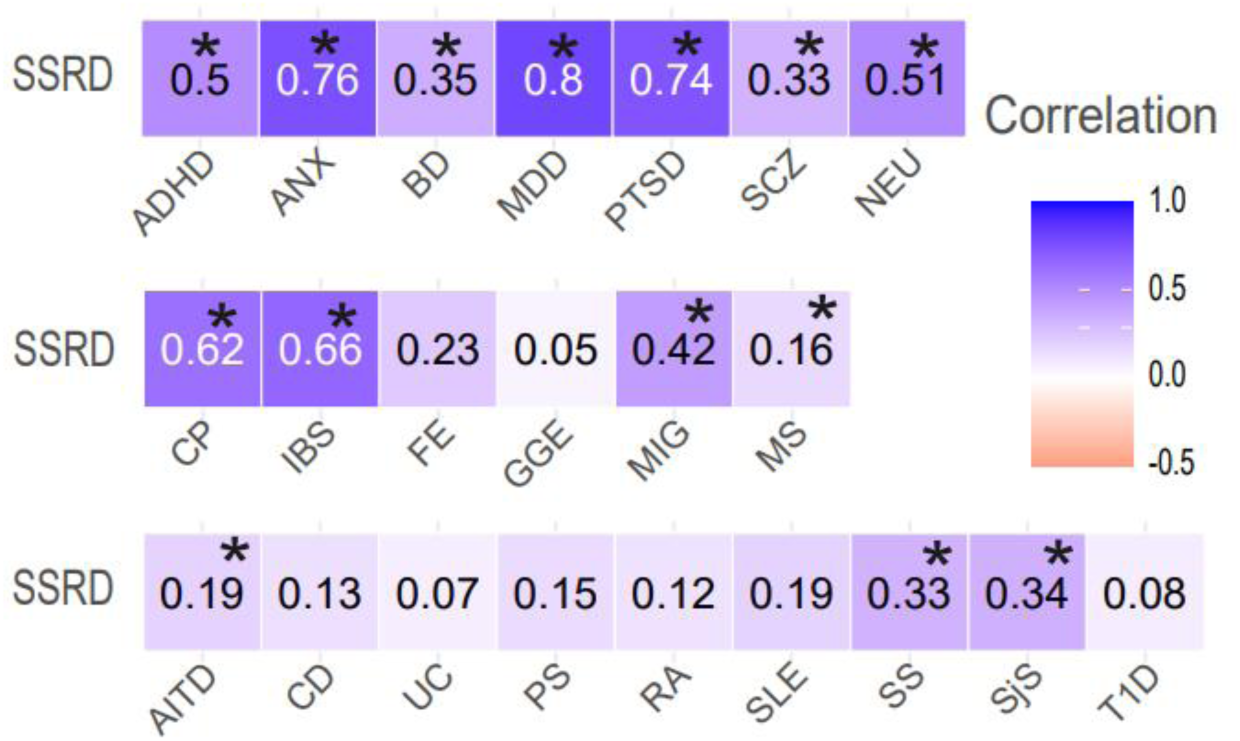
Genetic correlations between SSRD and psychiatric, neurological and immune-related phenotypes. Genetic correlations of SSRD with 22 phenotypes are shown: major psychiatric disorders (bipolar disorder ― BD, major depressive disorder ― MDD, schizophrenia ― SCZ), most-comorbid psychiatric disorders (anxiety ― ANX, attention deficit hyperactivity disorder ― ADHD, post-traumatic stress disorder ― PTSD), neurological disease (epilepsy, focal ― FE and generalised ― GGE, migraine ― MIG, multiple sclerosis ― MS), neuroticism ― NEU, chronic widespread pain ― CP (as a proxy of fibromyalgia), irritable bowel syndrome ― IBS, and immune-related disorders (psoriasis ― PS, type 1 diabetes ― T1D, systemic lupus erythematosus ― SLE, systemic sclerosis ― SS, Sjögren’s syndrome ― SjS, rheumatoid arthritis ― RA, autoimmune thyroid disorders ― AITD, Crohn’s disease ― CD and ulcerative colitis ― UC). For each phenotype, the genetic correlation (in numbers) is shown. Genetic correlations significant after Bonferroni correction (*p-*value <0.05/67) are marked with an asterisk. Detailed information is available in the Suppl. Table 3.

We performed a meta-analysis of SSRD, combining FND and SomD. One significant SNP was identified on chromosome 8 (rs7004685, Chr8:65565084) and mapped to the cytochrome P450 family 7 subfamily B member 1 gene (*CYP7B1)* and the basic helix-loop-helix family member e22 (*BHLHE22)* gene using the FUMA and OpenTarget approaches. In the subgroup SomD meta-analysis (n cases = 18,536, n controls = 1,819,203), two significant SNPs were identified, one lead SNP rs7817744, Chr8:65594917 (with a high LD – D value: 0.9781, R² 0.8977 – with rs7004685 for combined phenotype, mapped to the same genes, *CYP7B1* and *BHLHE22)* and second one rs2908882, Chr16:49074278 (see Table 2., Fig. 1, and Suppl. Fig. 4. for locus zoom plot). The gene mapped by the FUMA and the OpenTarget for rs2908882 was the cerebellin 1 precursor gene (*CBLN1)*. In the subgroup FND meta-analysis (n cases = 4,269, n controls = 1,852,274), no significant SNPs were identified (the details are described in the Suppl., Fig 3A and B.).

**Table 2.** Genomic loci associated with SSRD and SomD phenotypes (*p*-value < 5×10^−8^). Chr — chromosome number, BP — base pair position on build GRCh37 with start and stop end, lead SNP — lead single-nucleotide polymorphism, CADD score — Combined Annotation Dependent Depletion Score. *The SNPs on the CHR8 in the high LD between each other.

| Disorder | Chr | Lead BP: start-end, GRCh37 | Lead SNP | A1/A2 | P-value | CADD score | Z-score | Gene | Function |
| --- | --- | --- | --- | --- | --- | --- | --- | --- | --- |
| SSRD (FND& SomD) | 8 | 65565084: 65500797-65660906 | rs7004685* | A/G | 1.9e-10 | 20.7 | -6.4 | *CYP7B1*, *BHLHE22* | Protein coding |
| SomD | 8 | 65594917: 65500797-65608361 | rs7817744* | T/C | 1.1e-08 | 20.7 | -5.7 | *CYP7B1*, *BHLHE22* | Protein coding |
| SomD | 16 | 49074278: 49034109-49094077 | rs2908882 | T/C | 1.4e-08 |  | 5.7 | *CBLN1* | Protein coding |

LDSC analysis revealed a strong correlation between FND and SomD (rg=0.942, SE=0.11, *p=*3.85×10^-18^). The observed genetic correlation between FND and SomD did not differ significantly from 1 (*p*=0.299), supporting the combined meta-analysis and indicating a similar genetic background for both disorders. The observed-scale SNP heritability based on LDSC results for SSRD was 7.3% (SE=0.63), FND was 15.7% (SE=1) and SomD was 7.7% (SE=1). The SumHer (LDAK) tool revealed similar heritability for FND (was 15 %; SE=0.02), SomD (10 %; SE=0.01) and genetic correlation (rg=0.97, SE=0.08, p=1.83E-30) as LDSC.

We calculated the genetic correlation between SSRD, their subtypes, and psychiatric disorders and traits, neurological disorders, and immune-linked disorders (for the complete list of disorders see Fig. 2, Suppl. Fig. 5, and Suppl. Table 2 and 3). After Bonferroni correction for multiple testing, we found significant genetic correlation between SSRD and major psychiatric disorders (MDD, BD, SCZ), as well as anxiety, ADHD, and PTSD. Also, a significant correlation with migraine, IBS, and chronic pain was found. In addition, SSRD had significant correlation with autoimmune thyroiditis, multiple sclerosis, systemic scleroderma, and Sjögren’s syndrome (Fig. 2).

### MAGMA: gene-set and tissue expression enrichment analyses

MAGMA gene-set analysis did not reveal significantly enriched gene-sets or pathways (Suppl. Fig. 6) within all GWAS for the studied phenotypes.

### Validation analysis

In the validation cohort (Suppl. Table 4) two of the four genome-wide significant SNPs were in the same effect direction as the primary analyses. However, none of the identified loci were statistically significant, likely due to the small number of cases and controls in this cohort.

## Discussion

We have conducted the first GWAS meta-analysis on SSRD, identified the first genome-wide significant loci (chromosome 8), and one additional locus on chromosome 16, associated with SomD. We estimated the SNP-based heritability was 7.3 % and showed extensive genetic correlation with psychiatric, neurological, and immune-related diseases, as well as between the subgroups FND and SomD. Together, this indicates that genetics plays important role in the SSRD development, and SSRD has a common genetic background with psychiatric and neurological disorders.

For the SSRD meta-analysis, we found one significant SNP on chromosome 8 (rs7004685) that maps to the same genes (*CYP7B1* and *BHLHE22*) as for SomD analysis. The functional analysis revealed that *CYP7B1* is involved in the metabolism of neurosteroids, which play a crucial role in modulating synaptic activity, neuroplasticity, and neuroprotection [Stiles AR. et al., 2009; Yantsevich AV. et al., 2014]. Biallelic loss-of-function variants in *CYP7B1* were shown to cause hereditary spastic paraplegia type 5 [Prestsæter S. et al., 2020; Parodi L. et al., 2018; Schöls L. et al., 2017] and the SNP rs118111353 in *CYP7B1* is thought to be associated with Parkinson disease [Petkova-Kirova P. et al, 2024]. *CYP7B1* involvement in the metabolism of neurosteroids might be a possible link between the gene and functional symptoms as the influence of steroids in remodelling of the hippocampus and amygdala which regulate emotional reactivity and stress processing has been previously shown [Brown ES. et al., 1999; Fowler CH. et al., 2021].

The *BHLHE22* gene product belongs to the family of transcription factors and is expressed exclusively in the central nervous system and retina [Ge Y. et al., 2023]. Monoallelic and biallelic variants in this gene cause a neurodevelopmental disorder with agenesis of the corpus callosum, intellectual disability, and tone and movement abnormalities [Le C. et al., 2024]. Thus, we speculate that certain genetic variants in *BHLHE22* may affect development of cortical circuitry (especially those involving sensory, emotional, and motor processing and integration), predisposing individuals to functional disorders. Overall, our study provides new insights into potential genetic causes of functional disorders, with a possible biological connection to the current pathophysiological framework of these conditions.

In the SomD meta-analysis, one additional locus was revealed on chromosome 16: lead SNP rs2908882, the mapped gene *CBLN1*. The genetic variant rs2908882 has been identified in complex, non-specific psychiatric conditions [Kurki M. et al., 2023]. This locus was mapped to the *CBLN1,* which is critical for Purkinje cell synapse formation and maintenance. Additionally, *CBLN1* protein product influences synapse formation in cortical and hippocampal cells [McCormick LE. et al., 2022]. Given the cerebellum is believed to contribute to cognitive and emotional processing, *CBLN1*-dependent cerebellar dysfunction might hypothetically influence how the brain integrates sensory and emotional processing, leading to the development of functional symptoms. Moreover, according to the 2021 study by Gandhi PJ. et al., 2021, cerebellin 1 plays a crucial role in the mechanism of chronic pain. It is also strongly linked to the repetitive process underlying functional and somatic symptoms.

We found little support for previous findings of prior small-scale candidate gene studies. The tryptophan hydroxylase 2 (*TPH2*) gene [Spagnola P. et al., 2020] had a *p*-value of 0.18. The *TPH1* gene, rs1800532, and the oxytocin receptor *(OXTR)* gene, rs2254298 and rs53576 [Weber S. et al.; 2024, Jungilligens J. et al., 2022] were all non-significant (*p*-values 0.68, 0.53 and 0.32, respectively).

The estimated observed-scale SNP heritability was 7.3% for SSRD, and 15.7% for FND and 7.7% for SomD. These values are similar to the heritability estimates for psychiatric disorders such as anxiety disorders, ADHD or MDD, and neurological disorders such as migraine [Smeland OB. et al., 2024]. The level of comorbidity for psychiatric disorders in our study [see Suppl. 1., table 1] was in line with previous studies: the most common comorbid psychiatric disorder was MDD, generalized anxiety disorder, and PTSD. Previously, it has been reported that 51% [Thomas M. et al. 2006] to 95% [Feinstein A. et al., 2001; Patron VG. et al, 2023, Carle-Toulemonde G. et al., 2023] of adult FND patients have comorbid psychiatric conditions.

FND and SomD were highly genetically correlated (rg=0.94), suggesting a similar genetic susceptibility. Significant high genetic correlations were revealed for SSRD with psychiatric disorders (ADHD, anxiety disorders, BD, SCZ, MDD, PTSD) and neuroticism. Among psychiatric disorders, the correlation was particularly strong with the internalising disorders such as MDD, PTSD and anxiety. These results can support the existing theory of “shared polygenetic vulnerability” in brain-related disorders [Hindley G. et al, 2022; Greenwood TA. et al., 2020; Peterson RE. et al. 2021] and could explain high levels of comorbidity. The high correlation with neuroticism is also quite interesting: many studies implicate that psychopathological traits are highly pleiotropic and indicative for their largely interconnected nature crossing the borders of diagnostic classification [Grotzinger, AD. et al., 2019; Selzam et al., 2018]. The psychological studies also showed some common traits with personality traits: there are correlations with autistic traits [Ekanayake V. et al; 2017, McWilliams A et al., 2019; González-Herrero B. et al., 2023], alexithymia [Demartini B. et al., 2014] and altered emotional processing in people with FND [Pick S. et al., 2019]. A plausible hypothesis behind this significant correlation might be that the individuals with high scores for neuroticism are more likely to experience feelings of anxiety and fear, which can lead to an increased vulnerability to develop functional symptoms [Patron VG. et al, 2023], and our findings can support this psychological concept from a genetic perspective. Taken together, the genetic correlations of FND and SomD show similarities with the genetic continuum of neuropsychiatric disorders.

We demonstrated a significant genetic correlation between migraine and SSRD supporting previous findings [Bahrami S. et al., 2021; Smeland OB. et al, 2023; Brainstorm Consortium et al., 2018] about significant genetic correlation of migraine with different internalizing neuropsychiatric disorders (the correlation of migraine with depression was 0.32 which is in line with our data for SSRD) and high comorbidity level [Dresler T. et al., 2019]. This highlights the shared genetic architecture between migraine and SSRD, following their clinical overlap [McCracken HT. et al, 2024]. We also showed a significant correlation of SSRD with autoimmune disorders such as autoimmune thyroiditis, systemic scleroderma and primary Sjögren’s syndrome. This is in line with the recent paper of Joseph A. et al., 2024, which suggested a co-morbidity of FND with autoimmune disorders, and connective tissue disorders and endocrine disorders were more prevalent. These findings are in accordance with the previous studies which showed increased interleukin levels in SSRD patients [van der Feltz-Cornelis C. et al., 2023] as well in line with theory of the low-grade inflammation in functional somatic symptoms [McInnis PM. et al., 2020] which can activate stress system, fostering conditions that facilitate the onset of a functional disorder. This can also be supported by a highly significant genetic correlation between chronic pain and IBS, where the role of neuroinflammation in sensitization of chronic pain is well established [Ji RR. et al., 2018; Schumacher MA., 2024; Ishioh M. et al., 2024].

Our study has the strength of a large sample size, that enabled the discovery of the first GWAS significant loci for SSRD and the estimation of the heritability. We acknowledge that the diagnoses were collected from specialist health care diagnostics recorded in national health registries and electronic health records, which can be linked to a broader phenotype compared to prospective clinical assessments. However, for other brain-related diseases, the quality of data in these health registries is acceptable [Chen CY. et al., 2018; Smoller JV. et al., 2018]. Additionally, it was not possible to account for co-morbidities due to a small number of cases in each cohort. Therefore, we minimised this bias by using population controls in our GWAS design. However, we cannot rule out that SSRD comorbidities were excluded from all samples included in the large GWAS of neurological, psychiatric, and immune-related diseases, and thus affecting the genetic correlation results. We acknowledge that our focus was on individuals of European ancestry, mainly from Nordic populations, which may limit the generalizability of our findings.

In conclusion, we demonstrated substantial SNP heritability and identified novel disease-related loci for SSRD. We showed significant genetic correlation between the FND and SomD, and between SSRD and psychiatric disorders, migraine and some immune-related diseases. Our findings provide insights into the causes and molecular mechanisms of SSRD, highlighting underlying pathophysiological processes in SSRD and the overlap between diagnostic categories. Together, our findings can form the basis for the development of new therapies and the identification of disease biomarkers.

## Acknowledgements and Funding

Many thanks to our patients, who continue to inspire us to learn more about neurology and psychiatry through their complex conditions. We thank the International Parkinson and Movement Disorder Society (https://www.movementdisorders.org/) for their course on Neuropsychiatry in Movement Disorders, which motivated us to perform this analysis.

We thank the participants and researchers contributing to the GWAS investigated in the present study. The authors thank the researchers of the International MS Genetics Consortium (IMSGC) https://imsgc.net/ and consortia for access to summary-level data, as well as all participants who provided DNA samples. We want to acknowledge the participants and investigators of the FinnGen study: https://www.finngen.fi/ [Kurki M. et al., 2023].

The Norwegian Mother, Father and Child Cohort Study is supported by the Norwegian Ministry of Health and Care Services and the Ministry of Education and Research. We are grateful to all the participating families in Norway who take part in this ongoing cohort study. We thank the Norwegian Institute of Public Health (NIPH) for generating high-quality genomic data. This research is part of the HARVEST collaboration, supported by the Research Council of Norway (#229624). We also thank the NORMENT Centre for providing genotype data, funded by the Research Council of Norway (#223273), South-East Norway Version 7.0 3 Health Authorities, and Stiftelsen Kristian Gerhard Jebsen. We further thank the Center for Diabetes Research, the University of Bergen for providing genotype data and performing quality control and imputation of the data funded by the ERC AdG project SELECTionPREDISPOSED, Stiftelsen Kristian Gerhard Jebsen, Trond Mohn Foundation, the Research Council of Norway, the Novo Nordisk Foundation, the University of Bergen, and the Western Norway Health Authorities.

This research used data from the UK Biobank, a major biomedical database (https://www.ukbiobank.ac.uk/, accession number 27412).

We gratefully acknowledge *All of Us* participants for their contributions, without whom this research would not have been possible. We also thank the National Institutes of Health’s *All of Us* Research Program for making available the participant data examined in this study. The All of Us Research Program is supported by the National Institutes of Health, Office of the Director: Regional Medical Centers: 1 OT2 OD026549; 1 OT2 OD026554; 1 OT2 OD026557; 1 OT2 OD026556; 1 OT2 OD026550; 1 OT2 OD 026552; 1 OT2 OD026553; 1 OT2 OD026548; 1 OT2 OD026551; 1 OT2 OD026555; IAA #: AOD 16037; Federally Qualified Health Centers: HHSN 263201600085U; Data and Research Center: 5 U2C OD023196; Biobank: 1 U24 OD023121; The Participant Center: U24 OD023176; Participant Technology Systems Center: 1 U24 OD023163; Communications and Engagement: 3 OT2 OD023205; 3 OT2 OD023206; and Community Partners: 1 OT2 OD025277; 3 OT2 OD025315; 1 OT2 OD025337; 1 OT2 OD025276. In addition, the All of Us Research Program would not be possible without the partnership of its participants.

We gratefully acknowledge support from the Research Council of Norway (#223273, 296030, 324252, 326813, 334920, 344121), European Union’s Horizon 2020 research and innovation programme under the Marie Skłodowska-Curie Actions Grant 801133 (Scientia fellowship); the South-Eastern Norway Regional Health Authority (#2020060, IES), KG Jebsen Stiftelsen (SKGJ-MED-021). This project has received funding from the European Union’s Horizon 2020 research and innovation programme under grant agreements No 847776 and 964874, EEA grants (EEA-RO-NO-2018-0535, EEA-RO-NO-2018-0573), National Institutes of Health grants U24DA041123 and U24DA055330, NIH 5R01MH124839-02 (PGC4), and the Estonian Research Council (PRG1197). The EstBB part of the research was conducted using the Estonian Center of Genomics/Roadmap II funded by the Estonian Research Council (project number TT17).

We would like to thank all participants of the Estonian Biobank who have made this research possible, and the Estonian Biobank Research team – Andres Metspalu, Tõnu Esko, Reedik Mägi, Mait Metspalu, Mari Nelis and Georgi Hudjashov for data collection, genotyping, QC and imputation. Data analysis was carried out in part at the High-Performance Computing Center of the University of Tartu. Individual level data analysis in the Estonian Biobank was carried out under ethical approval 1.1-12/624 (24 March 2020) from the Estonian Committee on Bioethics and Human Research (Estonian Ministry of Social Affairs), using data according to release application 6-7/GI/8746 from the Estonian Biobank from the Estonian Biobank.

In the Swedish Schizophrenia Study the computations were enabled by resources provided by the Swedish National Infrastructure for Computing (SNIC) at Uppsala, partially funded by the Swedish Research Council through grant agreement no. 2018-05973.

We acknowledge the deCODE genetics/Amgen Inc team, Reykjavik, Iceland and Faculty of Medicine, University of Iceland, Reykjavik, Iceland: Ástrós Th. Skúladóttir, G. Bragi Walters, Thorgeir E. Thorgeirsson, Gyda Bjornsdottir, Hreinn Steffanson and Kari Stefansson for thoughtful comments and providing the data.

The acknowledgements for danish groups will be provide later.

This work was partly performed on the TSD (Tjeneste for Sensitive Data, projects NS9666S, NS9703S) facilities, owned by the University of Oslo and operated and developed by the TSD service group at the University of Oslo, IT Department (USIT), with resources for computation provided by UNINETT Sigma2 — the National Infrastructure for High Performance Computing and Data Storage in Norway.

## Data availability and Computational tools

GWAS data for meta-analysis will be available upon request, and in the GWAS catalogue after the one-year embargo following this publication.

**Analyses were performed using existing tools available on: Cleansumstat** GitHub_BioPsyk/cleansumstats

**FUMA** https://fuma.ctglab.nl/

**LDSC** https://github.com/bulik/ldsc **SumHer (LDAK)** https://dougspeed.com/ **MAGMA** https://cncr.nl/research/magma/ **METAL** https://github.com/statgen/METAL

**Python Convert** https://github.com/precimed/python_convert

## Abbreviation

ADHD: attention deficit hyperactivity disorder
*ADHFE1*: alcohol dehydrogenase iron containing 1 gene
AITD: autoimmune thyroiditis
ANX: anxiety
BD: bipolar disease
*BHLHE22*: basic helix-loop-helix family member e22
BP: base pair position
CADD score: combined annotation dependent depletion score.
CHR: chromosome number.
CNV: copy number variation
CHB: Copenhagen Hospital Biobank
FE: epilepsy, focal
FND: functional neurological disorders
FMD: functional movement disorders
DBDS: The Danish Blood Donor Study
*CBLN1*: cerebellin 1 Precursor gene
CHR: chromosome
CD: Crohn’s disease
CP: chronic widespread pain
*CYP7B1*: Cytochrome P450 family 7 subfamily B member 1 gene
IBS: irritable bowel syndrome
GGE: epilepsy, generalised
GWAS: genome-wide association study
LD: linkage disequilibrium
LDSC: linkage disequilibrium score regression
MDD: major depressive disorder
MHC: major histocompatibility complex
MIG: migraine
MOBA: Norwegian Mother, Father and Child Cohort Study
MRI: magnetic resonance imaging
MS: multiple sclerosis
NEU: neuroticism
*OXTR*: oxytocin receptor gene
PS: psoriasis
PTSD: post-traumatic stress-related disorder
RA: rheumatoid arthritis
SCZ: schizophrenia
SjS: primary Sjögren’s syndrome
SLE: systemic lupus erythematosus
SomD: somatoform disorders
SS: systemic sclerosis
SSRD: somatic symptom related disorders
Suppl.: Supplementary
T1D: type 1 diabetes
UC: ulcerative colitis
WES: whole exome sequencing
WGS: whole genome sequencing

