## Supplementary for "Genome-wide study of somatic symptom and related disorders identifies novel genomic loci and map genetic architecture"

**Material and methods, cohort description and GWAS:**

**MOBA cohort (Norway)**

The Norwegian Mother, Father and Child Cohort Study (MoBa) is a population-based pregnancy cohort study conducted by the Norwegian Institute of Public Health. Participants were recruited from all over Norway from 1999-2008. The women consented to participation in 41% of the pregnancies. The cohort includes approximately 114.500 children, 95.200 mothers and 75.200 fathers. The current study is based *on version 12* of the quality-assured data files released for research in 2019. The establishment of MoBa and initial data collection was based on a license from the Norwegian Data Protection Agency and approval from The Regional Committees for Medical and Health Research Ethics. The MoBa cohort is currently regulated by the Norwegian Health Registry Act. The current study was approved by The Regional Committees for Medical and Health Research Ethics (2016/1226/REK Sør-Øst C). We used the quality controlled genetic data from the MoBaPsychGen release v1 described by Corfield et al. [1]. The analysis was restricted to individuals of European genetic ancestry. More information about MoBa recruitment, questionnaires and data quality control are described elsewhere [2, 3]. We used Norwegian patient registry data (NPR 2008-2023) to define F44 and F45 patients with main diagnosis in all participants. The number of cases and controls available in Table 1, main text. The mean age F44 was 33 (SD=12.6), 83 % female. The mean age F45 36.2 (SD=11.4), 75 % female. For the exclusion of cases from the controls we used data from NPR, the same version, and *Kontroll og utbetaling av helserefusjoner* (KUHR, 2006-2023) database. Extraction of phenotypes and analysis in MOBA was performed in September 2024.

**UK Biobank cohort (the United Kingdom)**

This study used genotypes, demographic and clinical data from the UK Biobank, a large-scale biomedical database containing genotype and phenotype data for approximately 500,000 individuals [4]. Data for this study were obtained under accession number **27412.** The numbers of cases and controls available in table 1, main text. The mean age F44 was 55.4 (SD=8), 65 % female, and F45 was 56.4 (SD =8), 64 % female, at cohort recruitment. Genotyping, imputation, and central quality control procedures for the UK Biobank genotypes are described in detail elsewhere [5]. F44 and F45 cases were defined as we described in a main text: as a primary diagnosis at least one time in life, and controls were population controls without diagnosis F44/F45 elsewhere.

**Association testing**

The genome-wide association analysis was conducted using version 3 of the UK Biobank genetic data with the REGENIE v3.4.1 tool using Firth's logistic regression [6]. We selected White individuals as determined by self-declared ethnicity. Participants who had withdrawn their consent were removed. For the association analysis, we retained only autosomal variants with a minor allele count >20 and an imputation information score >0.8, leaving 19.9 million variants. Sex, age, and the first twenty genetic principal components were included as covariates.

**"All of Us" cohort (the USA)**

**Cohort description** NIH's *All of Us* Research Program is a longitudinal cohort study aimed at advancing precision medicine and improving human health through partnering with one million or more diverse participants across the USA. The information about cohort can be find here [7] and online https://www.researchallofus.org/.

For F44 cases we used cohort creation tool available through the workbench web interface defining filters: "Dissociative and conversion disorders_ICD_F44", "not Hispanic or Latino" participants, "White Race", chip-genotyped. For F45 cases we used enabled filters: "Somatoform_disorders_ICD_F45", "not Hispanic or Latino" participants, "White Race", genotype. In All of Us there were not possibility distinguish the main diagnosis from the secondary as well as to establish the source of diagnosis. For controls we used filters: "not Hispanic or Latino", "White Race", genotyped and excluded all samples included in cases. The mean year of birth in F44 cases was 1971 (SD = 17), 70 % female. The mean year of birth in F45 cases was 1964 (SD=16). 71 % female. In combined analysis for F44 and F45 the mean year of birth for cases was 1965 (SD=16). 71 % female.

**Genotyping and association testing**

Genotyping was described at All of Us Research Program Genomics Investigators, 2024 [8]. We work on the All of us data using All of Us Research Workbench under Individual Research Access. The analysis was based on the "ACAF threshold callset" which is derived from the short read WGS data restricted to variants that have a population-specific allele frequency greater than 1% or a population-specific allele count greater than 100 in any computed ancestry subpopulations (more details here https://support.researchallofus.org/hc/en-us/articles/4614687617556-How-the-All-of-Us-Genomic-data-are-organized). Using genetically inferred ancestry and relatedness as provided in the AllOfUs auxiliary resources for WGS data, we excluded individuals flagged as relatives as well as individuals with non-European genetic ancestry. This step resulted in n cases 469, n controls = 118174 (F44), n cases 1504, n controls 119219 (F45) and n cases = 1915, n controls = 116728 (F44 and F45) where association testing were performed. Association testing was performed using PLINK2.0 logistic regression https://www.cog-genomics.org/plink/2.0/formats#glm_logistic with sex, year of birth and 16 genetic principal components as covariates. Variants with minor allele frequency below 0.5% in the analysed sample were excluded from the association analysis.

**deCODE(Iceland)**

**Cohort description**

A large fraction of the Icelandic population has participated in a nationwide research program at deCODE genetics. Participants donated blood or buccal samples after signing a broad informed consent allowing the use of their samples and data in all projects at deCODE genetics approved by the National Bioethics Committee (NBC). Participants were genotypically verified as being of European descent [9]. The cases were defined using ICD-10 codes F44 and F45. Controls were all participants without any F44 or F45 (primary or secondary elsewhere in medical history) diagnoses. All personal identifiers of the participants' data were encrypted in accordance with the regulations of the Icelandic Data Protection Authority. The cases were identified from medical records, filed from 1987 to 2022, through collaboration with physicians at Landspitali—National University Hospital in Reykjavik, the Registry of Primary Health Care Contacts, and the Registry of Contacts with Medical Specialists in Private Practice. The data used in this study were approved by the NBC (VSNb201604001/03.01) following review by the Icelandic Data Protection Authority. For F44 GWAS N cases = 785 cases (72 % female) and 325403 controls. Mean Age 42.9 (SD=19.9). For F45 GWAS N cases = 2265 cases (68 % female) and 341147 controls. Mean Age 40.8 (SD=16.7). For F44+F45 N cases = 2931 cases (69 % female) and 341030 controls.

**Genotyping and imputation**

The genomes of 63,460 Icelanders were whole genome sequenced (WGS) [9, 10] using GAIIx, HiSeq, HiSeqX, and NovaSeq Illumina technology to a mean depth of 38×. Genotypes of

SNPs and indels were identified and called jointly with Graphtyper [11, 12]. Over 173,000 Icelanders were genotyped using various Illumina SNP arrays [9, 10]. The genotypes were long-range phased [13] to improve genotype calls using haplotype sharing information. Subsequently, extensive encrypted genealogic information was used to impute variants into the chip-typed Icelanders, as well as ungenotyped close relatives [14] to increase the sample size and power for association analysis.

**Association testing**

We applied logistic regression, assuming an additive model, using the three traits as the dependent variable and the genotype counts as the independent variable. Likelihood ratio test was used to compute P-values. The variants included in the analysis had imputation information above 0.8 and MAF over 1%. We adjusted for sex, county of origin, age at data analysis or age at death (first and second order terms included), blood sample availability for the individual, and an indicator function for the overlap of the lifetime of the individual with the time span of phenotype collection. We used LD score regression to account for distribution inflation due to cryptic relatedness and population stratification [15] and used the intercepts as correction factors ($CF_{F44}$ = 1.04, $CF_{F45}$ = 1.08, $CF_{F44+F45}$ = 1.09). For association testing we used deCODE software developed at deCODE genetics [9, 14].

**Estonian Biobank (Estonia)**

**Cohort description:**

The Estonian Biobank (EstBB) is a population-based biobank with over 210,000 individuals (about 20% of Estonia's adult population). The participants had to join the biobank voluntarily and be at least 18 years old. All individuals have signed a broad consent form and completed a health- and lifestyle-related questionnaire upon recruitment. All activities at the EstBB are regulated by the Estonian Human Genes Research Act**.** Detailed information about diagnoses and prescriptions is obtained by regularly linking the EstBB database with the national health insurance fund database and other relevant databases (national health registries, hospital databases). The diagnoses are recorded according to ICD-10 codes [16].

The analysis was conducted using the data freeze, which contained 211,658 individuals who joined EstBB between 2002 and 2023 (February) and were born between 1905-2005 (median 1973). Individual level data analysis in the Estonian Biobank was carried out under ethical approval 1.1-12/624 (24 March 2020) from the Estonian Committee on Bioethics and Human Research (Estonian Ministry of Social Affairs), using data according to release application 6-7/GI/8746 from the Estonian Biobank. The proportion of females was 65.4%. Among the participants, 98.7% were of European ancestry, 1.1% were unknown (data unavailable) and 0.2% of other ancestry. For the genome-wide study, cases were defined as participants with the ICD-10 codes F44* or F45* as a main diagnosis and diagnosed by a neurologist or psychiatrist. This information was received from the Estonian Health Insurance Fund (data from 2004 until 2022). The controls were all undiagnosed biobank participants without these specific ICD-10 codes anywhere in the medical records including self-reported diagnoses. After quality control was conducted on phenotype data (duplicated samples and individuals with less than five different ICD-10 codes were removed) and genotype data (participants with non-European ancestry, without PCs or genotype data were excluded) we conducted three GWASes with the following case-control numbers: F44* (n_cases=506, n_controls=205,244), F45* (n_cases=4,530, n_controls=173,222) and F44*-F45* (n_cases=4,960, n_controls=172,672).

**Genotyping and imputation**

Estonian Biobank samples were genotyped at the Core Genotyping Lab at the Institute of Genomics, University of Tartu, using Illumina GSAv1.0, GSAv2.0, GSAv2.0_EST, and GSAv3.0_EST arrays. Genotype data quality control was performed according to best practices (individuals with call rate <95%, who deviated ±3SD from the samples' heterozygosity rate mean or showed mismatch between heterozygosity of the X chromosome and sex based on phenotype data were excluded; all AT and GC SNPs, invariable SNPs, SNPs showing potential traces of batch bias, poor cluster separation results and inconsistent allele frequency among any of the EstBB genotyping experiments were removed). Pre-phasing was conducted with Eagle v2.4.1 [17]. For imputation, the population-specific hg38 imputation reference panel of 2,695 WGS samples was used [18]. Phased genotyping data was first lifted over to hg38 coordinates using Picard liftover (LiftoverVcf v2.26, file hg19ToHg38.over.chain) and imputation was done using Beagle 5.4 (version: 22Jul22.46e) [19].

**Association testing**

The GWAS was conducted with REGENIE v3.2 using approximate Firth logistic regression6 with sex, year of birth and first ten genetic principal components as covariates.

Computations were carried out in the High Performance Computing Center of the University of Tartu.

**Copenhagen Hospital Biobank (CHB) and The Danish Blood Donor Study (DBDS) (Denmark)**

**Cohort description**

Cases with F44/F45 primary diagnosis were selected from Copenhagen hospital biobank (CHB) and the Danish Blood donor study (DBDS, 20). Controls are selected from DBDS. CHB is an ongoing biobank that includes surplus material from diagnostic testing on individuals admitted at hospitals in the Danish Capital Region. It has been incorporating patients from The Copenhagen University Hospital (Rigshospitalet) since 2009 and was expanded in 2012 to include patients from the entire Capital Region. There are no exclusion criteria. Genotyping of CHB samples was performed at deCODE Genetics, Iceland under the CHB study on pain and degenerative musculoskeletal diseases (CHB-PDS) with approval number: NVK-1803812, P-2019-51. The gender distribution of cases from CHB-PDS comprises 70.39% women and 29.61% men, and the participants are aged between 23 and 102 years. The mean ages for women and men are 57 (23.53 IQR) and 55 (31 IQR), respectively. The sample is based on European ancestry.

The Danish Blood Donor Study (DBDS) is an ongoing nationwide research initiative established in 2010 to investigate the impact of blood donation on health. By 2015, it encompassed all blood donation facilities in Denmark. Participants provide a whole-blood sample for DNA extraction and complete several rounds of questionnaires concerning their health, social factors, and lifestyle. The gender distribution of the DBDS is approximately equal, comprising 49.53% women and 50.47% men, and includes participants aged 19 to 87 years. The mean ages for women and men are 50 (21.53 IQR) and 48 (22.7 IQR), respectively [21, 22]. The sample is based on European ancestry. The genetic study under DBDS has approval number NVK-1700407, P-2019-99.

**Genotyping and imputation**

The samples were genotyped on two versions of the Human Global Screening Arrays (GSA) (v1.0 and v3.0). Phasing was conducted using Eagle software. Imputation relied on a reference set of whole genome sequenced North-Western Europeans, which was called using GraphTyper [11, 12]. The deCODE in-house developed imputation workflow was employed for imputation [14]. The genetic data is in build hg38. Imputation is carried out at deCODE genetics following internal QC [14]. A joint variant calling with GraphTyper [17] forms the basis of the imputation. Before imputation, duplicate samples, samples failing the sex check, those with a genotype rate of less than 0.98, and ancestry outliers were removed [23] using EIGENSOFT (v 6.0.1) [24]. Data was also filtered for a genotype rate higher than 0.95, variants with a minor allele frequency (MAF) exceeding 0.01, variants with a P-value < 10^(-6) for Hardy-Weinberg equilibrium, and LD pruning using a window size of 100 markers that shifted by 25 markers, removing half of every variant pair with a genotypic r^2 > 0.1. The LD-pruned markers were utilised to calculate heterozygosity, identity by state, and sex; the following samples were removed: those with outlying heterozygosity (5 > SD from the median), one sample from each pair of samples with an IBS > 0.9, and all samples where reported sex did not match that determined from genotypes. Finally, all A/T and C/G markers were removed to avoid strand issues.

**Association testing**

Cases were defined in CHB and DBDS while controls were defined in DBDS. The analysis was conducted using fastGWA with the --fastGWA-mlm-binary option, utilising a generalised linear mixed model (GLMM-) based GWAS analysis for binary traits, furthermore, adjusting for sex, age, batch, and the first twenty genetic principal components as covariates.

**FinnGen (Finland)**

The FinnGen study is a large-scale genomics initiative that has analyzed over 500,000 Finnish biobank samples and correlated genetic variation with health data to understand disease mechanisms and predispositions [25]. The project is a collaboration between research organizations and biobanks within Finland and international industry partners. We used summary statistics publicly available from R12 release (Public release: November 4, 2024) of FinnGen. N cases F44 1525 F5_DISSOCIATIVE, 82 % females, median age at the first event (years) 32. N cases F 45 7148, F5_SOMATOFORM, 71 % females, median age at the first event (years) 46.

**Sweden Schizophrenia Study (Sweden)**

For replication analysis, we used samples from the Sweden Schizophrenia Study [26-30]. Dried blood spot samples were collected from the neonatal screening biobank (PKU biobank).

Participants were identified by the National Board of Health and Welfare based on being born 1981-1999, with information on birth hospital and known mother in the medical birth register. Randomly selected individuals born in Sweden 1981-1999 and their biological mothers were identified in the MBR for controls. All procedures were in accordance with an approval from the Stockholm regional review board (Dnr. 2016/1130-31/2). Individual consent was not required. Neonatal dried blood spots, originally collected for the purpose of the National Screening Program at Centrum för Medfödda Metabola Störningar, Karolinska University Hospital (Solna, Sweden) have been stored in a controlled environment since 1981. Two 3-mm diameter discs were retrieved from each blood spot, placed in a 96-well plate, with proteins eluted on a rotary shaker. Plates were then stored at -80°C and shipped on dry-ice to Statens Serum Institut (Denmark) for genotyping in 2020-22. The same experimental approach was used as previous studies [31]. 200 µL DNA was extracted using Extract-N-Amp Blood PCR Kit (Sigma-Aldrich), amplified in triplicate using the REPLI-g kit (Qiagen) and then combined. Amplified DNA was genotyped using the Illumina GSA v3 +MD according to manufacturing directions, with the exception that 7µL DNA was denatured in 1µL 0.4N NaOH. Cases with F44 or F45 were identified using the Swedish inpatient and outpatient discharge register, which captures all outpatient and inpatient hospitalizations [32, 33] since 1987. The register contains ICD-10 discharge diagnoses made by attending physicians for each hospitalization. Case inclusion criteria included ≥1 hospitalizations with a discharge diagnosis of either F44 or F45. Controls were selected from Swedish population registers and included no contacts for schizophrenia or schizoaffective disorder. N cases F44=87, N controls=2239, N cases F45=72, N controls=2239. F44 or F45 N cases: 159, N controls: 2239.

**Association analysis**

Quality control of the genotype was performed according to the RICOPILI pipeline [34]. This included SNP missingness <0.05 (before removing samples); sample missingness <0.02; |Fhet| <0.2; SNP missingness <0.02 (after removing samples); minor allele frequency >0.01; difference in SNP missingness between cases and controls <0.02; and SNP Hardy–Weinberg equilibrium (P>10−6 in controls or cases). When case-control designation is mentioned, it was regarding schizophrenia case/control status. Imputation was performed using the Haplotype Reference Consortium (r1.1) on the Sanger Imputation Server. Pairs of participants with relatedness (KINSHIP value: MZ twin >0.354, 1st-degree relative: >0.25, 2nd-degree relative:

>0.125) were identified and then a sample was removed to retain cases (if case-control) or the sample with higher call rate (if case-case or control-control).

We conducted PCA for batches combined with the 1000 Genomes Project genotypes (Five major global populations included as the ancestry reference) using directly genotyped overlapping SNPs using the following [34]. Ancestry outliers were removed if they were >3SD from the average PC1 or PC2 for the European reference. Upon removing the ancestry outliers, the genetic ancestry PCs were re-computed on imputed genotypes and used in the association as covariates.

We performed association testing with case/control status using an additive logistic regression model in PLINK2, including the first five genetic ancestry principal components. We conducted 3 GWAS': (1) F44, (2) F45, or (3) F44 or F45.

**Fig. 1.** Results of PubMed search for different disorders AND genetics (September 2024). We see a significant underrepresentation of the numbers of manuscripts linked to functional neurological and somatoform disorders (FND/SomD) compared to other neurological and psychiatric disorders. *FND was requested as (("FND" OR "Functional Neurological Disorders" OR "Non-epileptic Seizures" OR "Psychogenic Movement Disorders" OR "Psychogenic Seizures" OR "Psychogenic Disorders" OR "Dissociative Disorders" OR "Conversion Disorders" OR "Functional Seizures") AND ("Genetics") according to recent changes in classification, and only 10 papers among 187 (5.3%) were research articles that study genetics in FND (usually association testing of one SNP but not a genome-wide association studies).

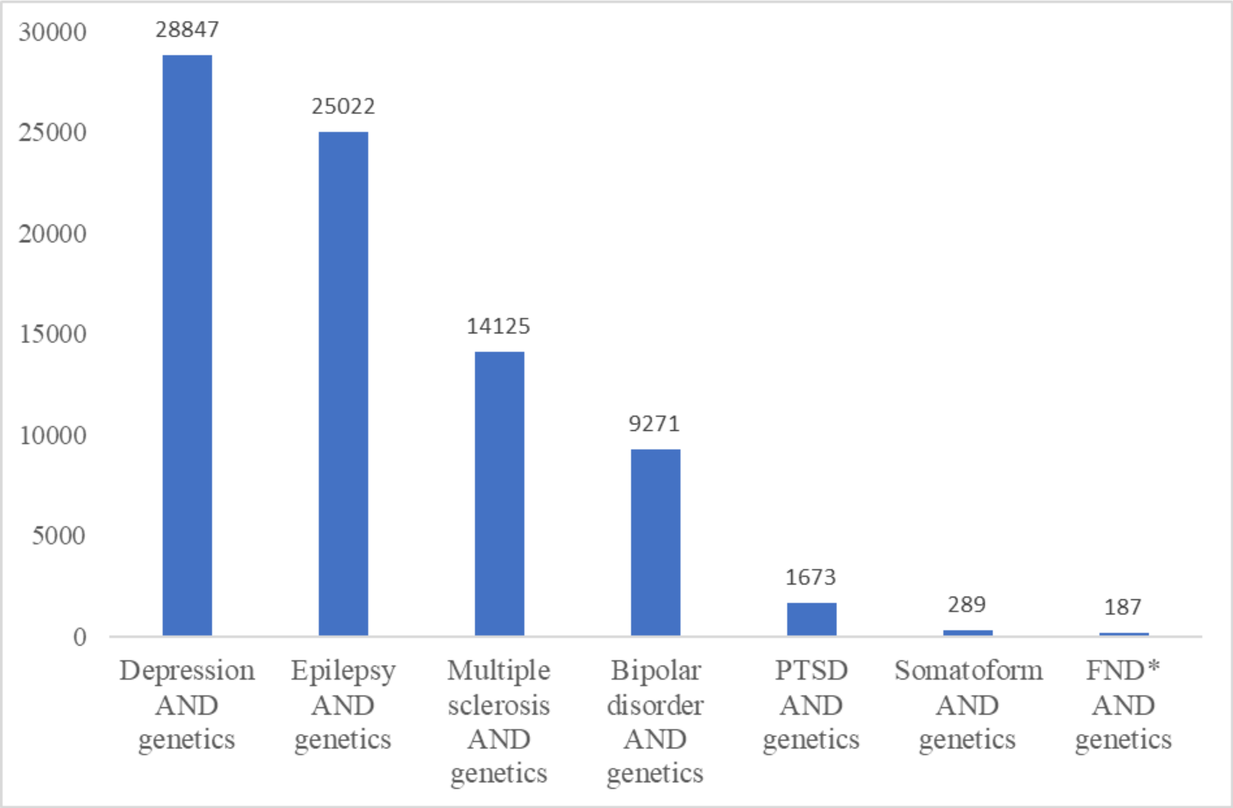

**Fig.2. QQ plots for GWAS meta-analyses (FND F44, SomD F45, SSRD F44 and F45).**

**A. FND (F44).**

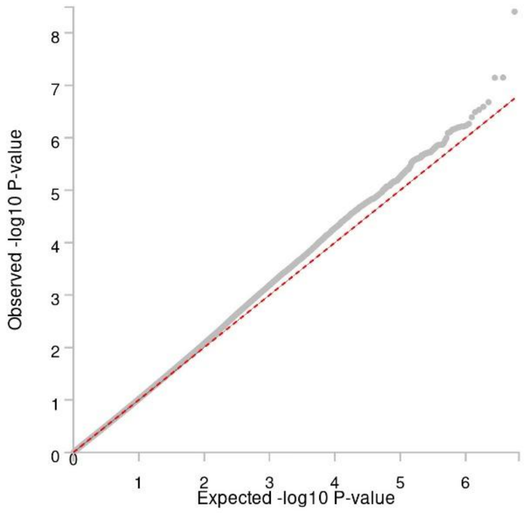


**B. SomD (F45).**

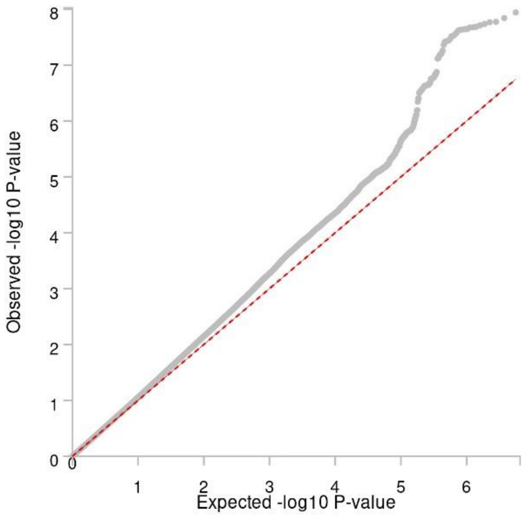


**C. SSRD (F44 and F45).**

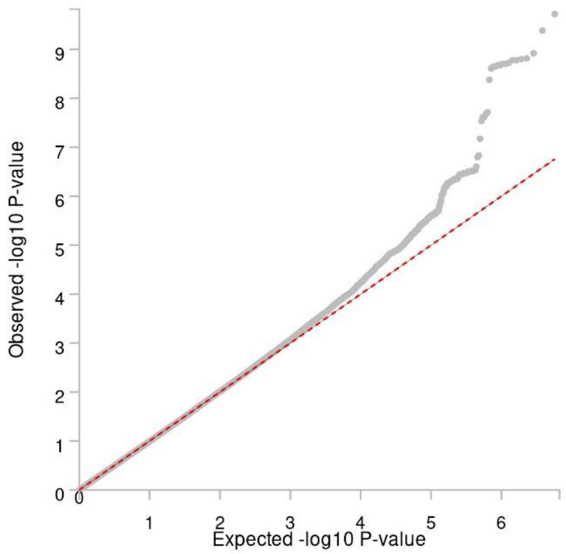

**Fig. 3. A. Manhattan plot for the discovery genome-wide association analysis in FND (blue plot).** −log10 *p*-values are plotted for all variants across chromosomes 1–22. The bold dots indicate loci with a lead variant genome-wide association study *p*-value of < $5\times10^{-8}$. The gray line indicates the threshold for genome-wide significance (*p*-value of $5\times10^{-8}$). **B.** Locus zoom plot for rs7830497.

In the FND meta-analysis (n cases = 4,269, n controls = 1,852,274, n markers = 14,236,292), no significant SNPs supported by tower on Manhattan plot were identified. However, we identified one significant SNP, rs7830497 (Chr8:67226413, see Supplementary Table 2. and Fig. 3 A and B). The mapped genes defined by FUMA were alcohol dehydrogenase iron containing 1 gene (*ADHFE1)* and *C8orf46*. The OpenTarget algorithm also mapped this locus to the *ADHFE1* gene. For F44 GWAS we performed the analysis of SNPs in LD with the leading SNP and the nearest SNP in the current GWAS had a p-value $7.2\times10{-}6$ (rs62511213, r2 = 0.832). This possibly explains why there is no tall stack on chromosome 8 under the significant SNP. However, based on minor allele frequency for rs7830497 for all cohorts < 1 % and lack of the tall stack on the Manhattan plot we need to highlight that this finding should be treated with caution.

**A. Manhattan plot**

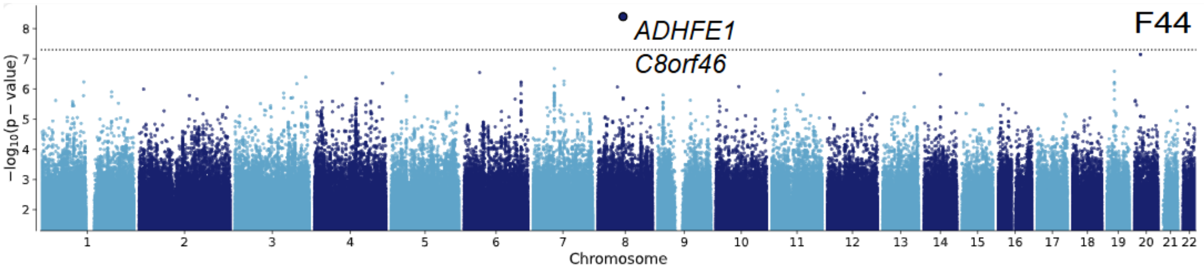


**B. Locus zoom plot.**

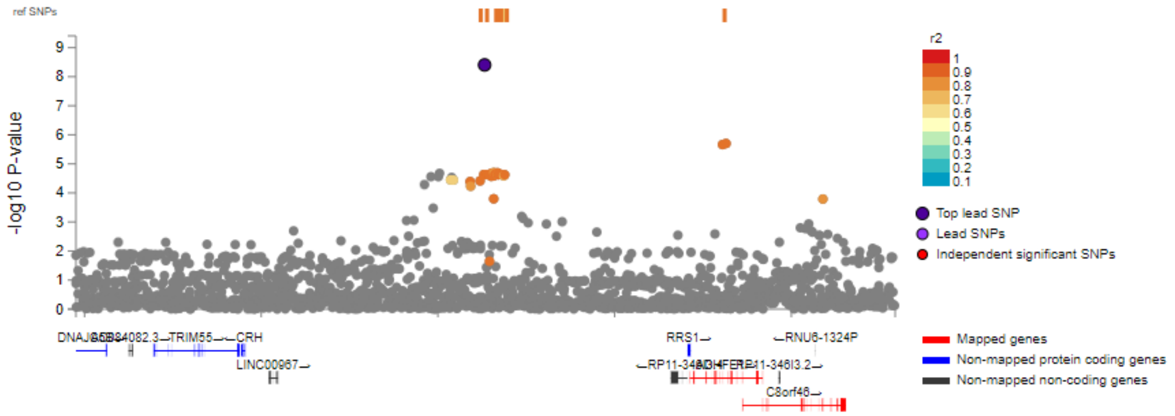

**Fig. 4. Locus zoom plot.**

A. SomD (F45) CHR8. rs7817744

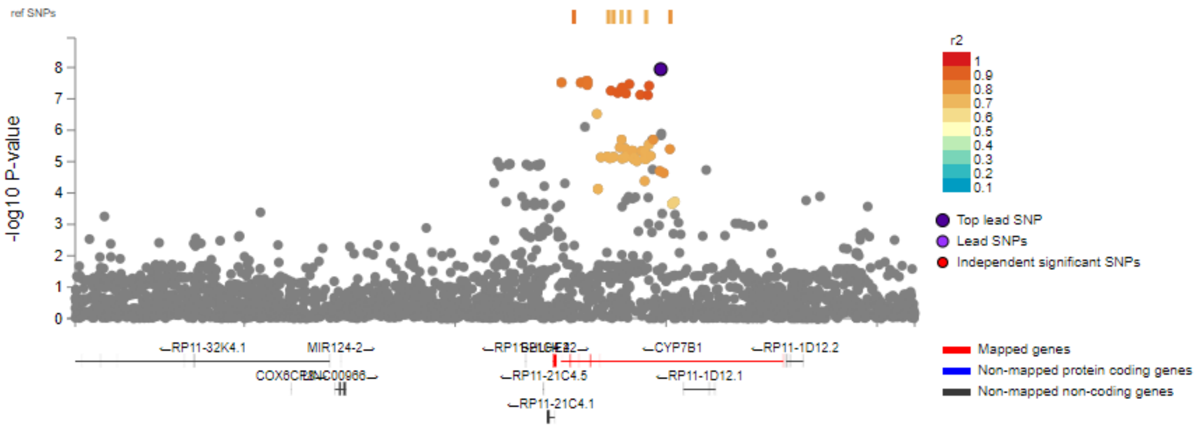


B. SomD (F45) CHR16. rs2908882.

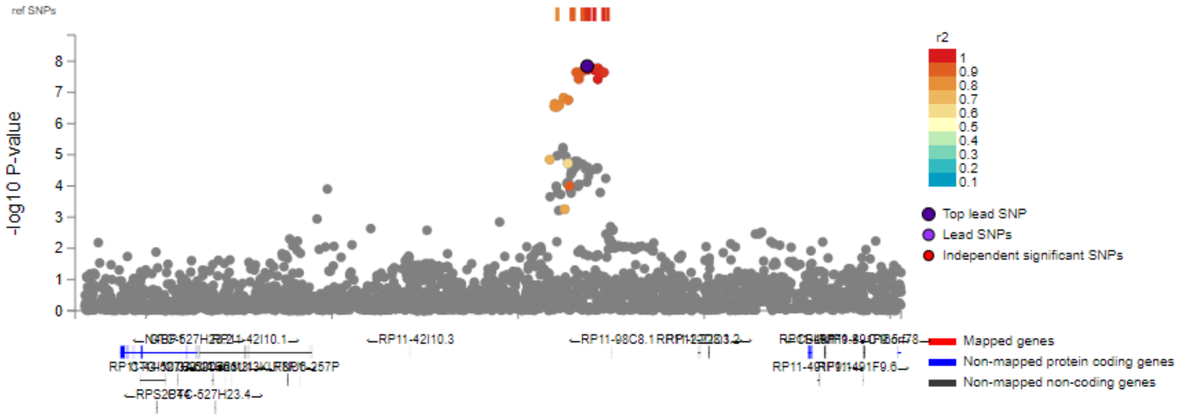


C. SSRD (F44 and F45) rs7004685.

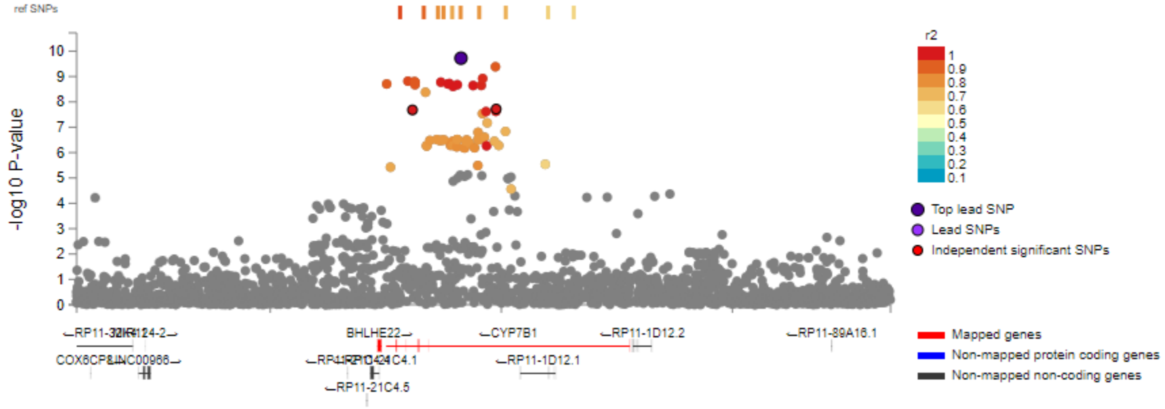

**Fig. 5. Genetic correlations between functional neurological disorders, somatoform disorders and other phenotypes.**

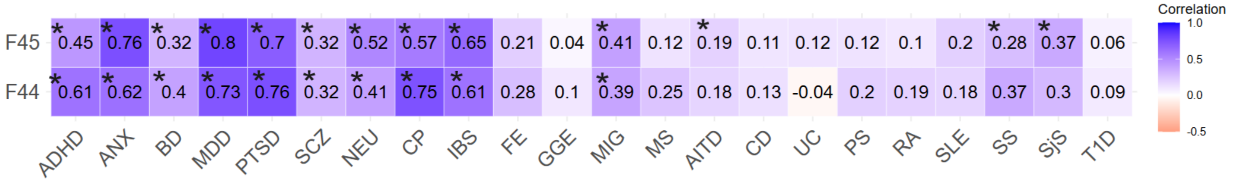


Genetic correlations of study traits with 22 phenotypes were shown: major psychiatric disorders (bipolar disease — BD, major depressive disorder — MDD, schizophrenia — SCZ), most-comorbid psychiatric disorders (anxiety — ANX, attention deficit hyperactivity disorder — ADHD, post-traumatic stress disorder — PTSD), neurological disease (epilepsy, focal — FE and generalised — GGE, migraine — MIG, multiple sclerosis — MS), neuroticism — NEU, chronic widespread pain — CP (as a proxy of fibromyalgia), irritable bowel syndrome — IBS, and immune linked disorders (psoriasis — PS, type 1 diabetes — T1D, systemic lupus erythematosus — SLE, systemic sclerosis — SS, Sjogren's syndrome — SjS, rheumatoid arthritis — RA, autoimmune thyroid disorders — AITD, Crohn's disease — CD and ulcerative colitis — UC). For each phenotype, the genetic correlation (in numbers) is shown. Genetic correlations significant after Bonferroni correction (p-value <0.05/67, taking into account combined SSRD in the main text) are marked with an asterisk.

**Fig.6. MAGMA results: GTEx v8 54 tissue types.**

**A. FND (F44).**

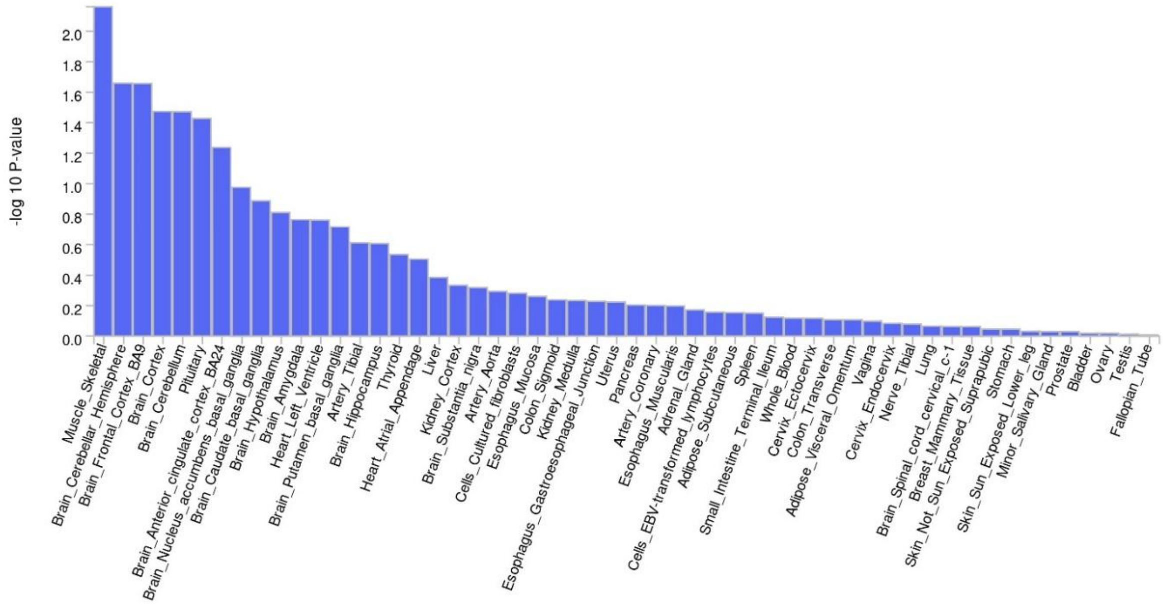

**B. SomD (F45).**

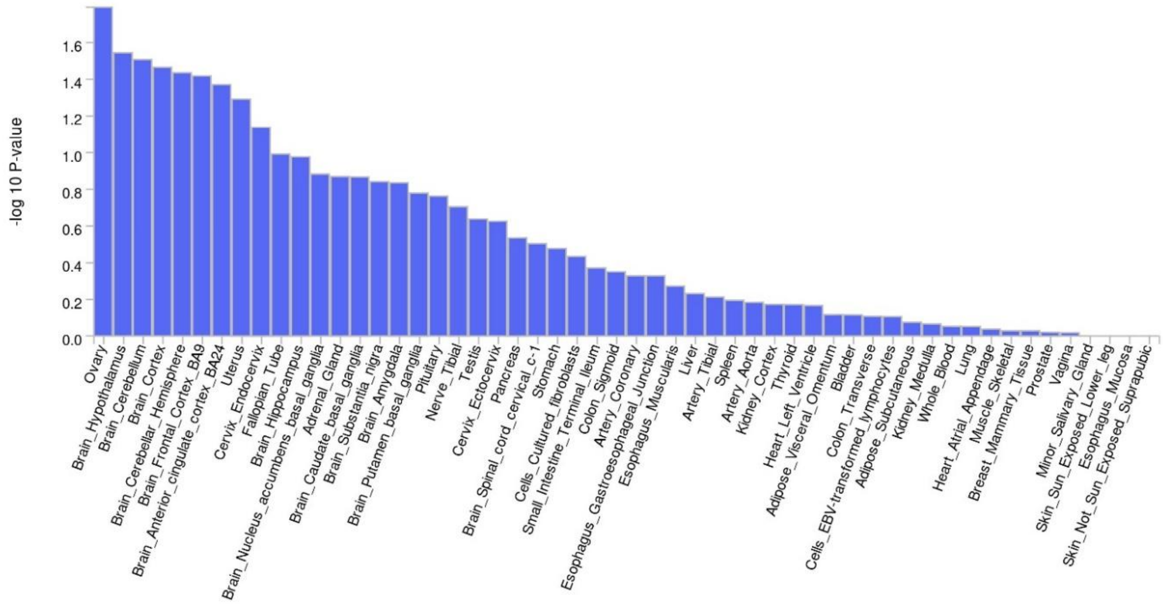


**C. SSRD (F44 and F45).**

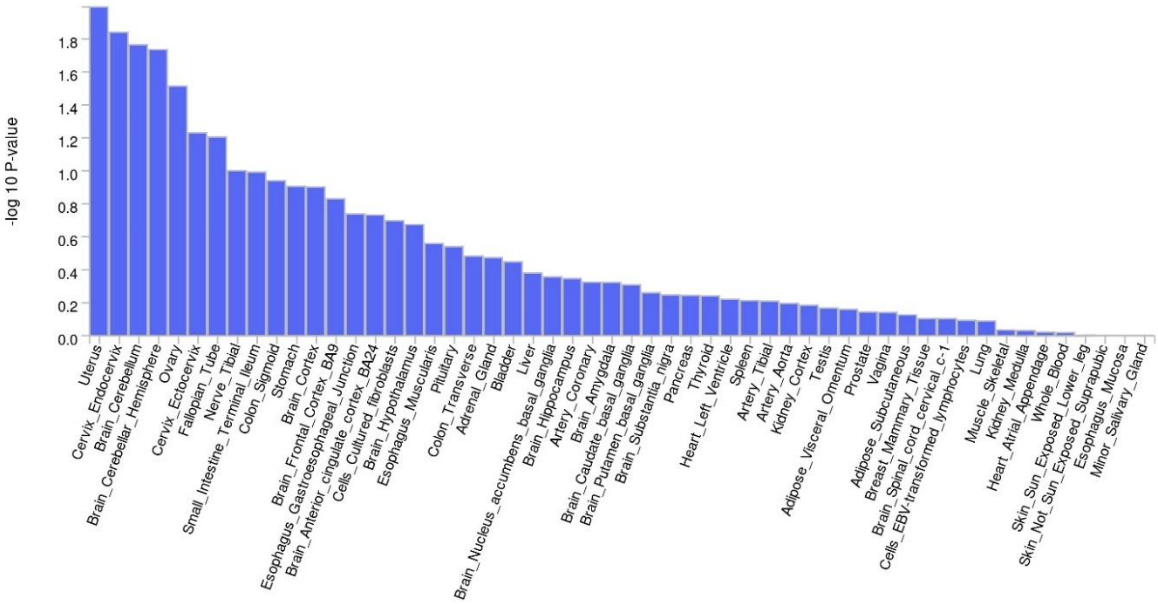

### Supplementary Table 1. Top co-morbid 5 diagnoses*:

**Iceland**

| **F44** | **ICD 10 code** | **Name ICD 10 code** | **F44 comorbidity, percent** |
|---|---|---|---|
| | F41 | Other anxiety disorders | 45 |
| | F32 | Depressive episode | 37 |
| | F43 | Reaction to severe stress, and adjustment disorders | 25 |
| | F33 | Recurrent depressive disorder | 20 |
| | F10 | Mental and behavioral disorders due to use of alcohol | 19 |
| **F45** | **ICD 10 code** | **Name ICD 10 code** | **F45 comorbidity, percent** |
| | F41 | Other anxiety disorders | 42 |
| | F32 | Depressive episode | 32 |
| | F33 | Recurrent depressive disorder | 20 |
| | F17 | Mental and behavioral disorders due to use of tobacco | 19 |
| | F10 | Mental and behavioral disorders due to use of alcohol | 19 |

**Estonia**

| **F44** | **ICD 10 code** | **Name ICD 10 code** | **F44 comorbidity, percent** |
|---|---|---|---|
| | F32 | Depressive episode | 55 |
| | F41 | Other anxiety disorders | 54 |
| | F45 | Somatoform disorders | 39 |
| | F33 | Recurrent depressive disorder | 34 |
| | F43 | Reaction to severe stress, and adjustment disorders | 33 |
| **F45** | **ICD 10 code** | **Name ICD 10 code** | **F45 comorbidity, percent** |
| | F41 | Other anxiety disorders | 56 |
| | F32 | Depressive episode | 52 |
| | F33 | Recurrent depressive disorder | 27 |
| | F43 | Reaction to severe stress, and adjustment disorders | 23 |
| | F48 | Other neurotic disorders | 17 |

**Norway**

| **F44** | **ICD 10 code** | **Name ICD 10 code** | **F44 comorbidity, percent** |
|---|---|---|---|
| | F43 | Reaction to severe stress, and adjustment disorders | 51 |
| | F32 | Depressive episode | 36 |
| | F41 | Other anxiety disorders | 31 |
| | F33 | Recurrent depressive disorder | 24 |
| | F40 | Phobic anxiety disorders | 19 |
| **F45** | **ICD 10 code** | **Name ICD 10 code** | **F45 comorbidity, percent** |
| | F41 | Other anxiety disorders | 23 |
| | F43 | Reaction to severe stress, and adjustment disorders | 22 |
| | F32 | Depressive episode | 19 |

| | | | |
|---|---|---|---|
| | F33 | Recurrent depressive disorder | 12 |
| | F40 | Phobic anxiety disorders | 11 |

**UKB**

| F44 | ICD 10 code | Name ICD 10 code | F44 comorbidity, percent |
|---|---|---|---|
| | F32 | Depressive episode | 66 |
| | F41 | Other anxiety disorders | 65 |
| | F17 | Mental and behavioural disorders due to use of tobacco | 18 |
| | F05 | Delirium unspecified | 13 |
| | F43 | Reaction to severe stress, and adjustment disorders | 8 |
| **F45** | **ICD 10 code** | **Name ICD 10 code** | **F45 comorbidity, percent** |
| | F41 | Other anxiety disorders | 19 |
| | F32 | Depressive episode | 17 |
| | F17 | Mental and behavioural disorders due to use of tobacco | 7 |
| | F10 | **Mental and behavioural disorders due to use of alcohol** | 5 |
| | F05 | Delirium unspecified | 2 |

*These data were not available for Denmark, AllofUs, FinnGen.

| Supplementary Table 2. GWAS used for LDSC correlation. | | | | | | | | |
|---|---|---|---|---|---|---|---|---|
| | GWAS available | Abbreviation | N cases | N controls | N effective | Year | Source | Availability |
| | **Psychiatric phenotypes** | | | | | | | |
| 1 | Bipolar Disease | BD | 59287 | 781022 | 150670 | 2025 | PMID 39843750 | Publicly available |
| 2 | Major Depressive Disorder | MDD | 412305 | 1588397 | 1152651 | 2025 | PMID 39814019 | Publicly available |
| 3 | Schizophrenia | SCZ | 53386 | 77258 | 126282 | 2022 | PMID 35396580 | Publicly available |
| 4 | Attention deficit hyperactivity disorder | ADHD | 38691 | 186843 | 128214 | 2023 | PMID 36702997 | Publicly available |
| 5 | Anxiety | ANX | 31977 | 82144 | 92068 | 2020 | PMID 31748690 | Publicly available |
| 6 | Post-traumatic stress-related disorder | PTSD | 137136 | 1085746 | 641533 | 2024 | PMID 38637617 | Publicly available |
| | **Neurological phenotypes** | | | | | | | |
| 7 | Epilepsy, focal | FE | 14939 | 42436 | 44197 | 2023 | PMID 37653029 | Publicly available |
| 8 | Epilepsy, generalized | GGE | 6952 | 42436 | 23894 | 2023 | PMID 37653029 | Publicly available |
| 9 | Migraine | MIG | 74495 | 1259808 | 281344 | 2023 | PMID 37884687 | Publicly available |
| | **Immune disorders** | | | | | | | |
| 10 | Multiple sclerosis | MS | 14802 | 26703 | 38093 | 2019 | PMID 31604244 | Available by request, IMSGC |
| 11 | Psoriasis | PS | 17255 | 693100 | 64320 | 2023, 2021, 2012 | R9 FinnGen, PMID 34278373, PMID 22482804 | In house meta-analysis from publicly available data [35] |
| 12 | Rheumatoid Arthritis | RA | 22350 | 74823 | 68838 | 2022 | GCST 90132223 | Publicly available |
| 13 | Systemic Sclerosis | SS | 9095 | 17584 | 23978 | 2019 | GCST 31672989 | Publicly available |
| 14 | Primary Sjogren's syndrome | SjS | 3232 | 17481 | 10911 | 2022 | PMID 35896530 | Publicly available |
| 15 | Systemic Lupus Erythematosus | SLE | 5595 | 361571 | 15096 | 2015 | R9 FinnGene, PMID 26502338, PMID 29848360 | In house meta-analysis from publicly available data [35] |
| 16 | Diabetes type 1 | T1D | 18,942 | 501,638 | 73011 | 2021 | PMID 34012112 | Publicly available |
| 17 | Autoimmune thyroid disorders | AITD | 30234 | 725172 | 114296 | 2020 | PMID 32581359 | Publicly available |
| 18 | Crohn's Disease | CD | 12194 | 34915 | 36151 | 2017 | PMID 28067908 | Publicly available |

| | | | | | | | | |
|---|---|---|---|---|---|---|---|---|
| 19 | Ulcerative Colitis | UC | 12366 | 34915 | 36527 | 2017 | PMID 28067908 | Publicly available |
| | **Borderline personality phenotype** | | | | | | | |
| 20 | Neuroticism | NEU | 390278 | * total sample size | | 2018 | PMID 29942085 | Publicly available |
| | **Mixed phenotypes** | | | | | | | |
| 21 | Irritable bowel syndrome | IBS | 40548 | 293220 | 142488 | 2021 | PMID 34741163 | Publicly available |
| 22 | Chronic widespread pain (proxy of fibromyalgia) | CP | 6021 | 427884 | 23750 | 2023 | GCST90129441 | Publicly available |

**Supplementary Table 3. LDSC analysis for correlation between FND, SomD and psychiatric and immune disorders**

| | rg | p-value | rg | p-value | rg | p-value | rg | p-value |
|---|---|---|---|---|---|---|---|---|
| | **ANX** | | **MDD** | | **PTSD** | | **SCZ** | |
| **F44** | 0.617 | 1.82E-12 | 0.725 | 8.64E-28 | 0.757 | 1.81E-27 | 0.323 | 2.11E-08 |
| **F45** | 0.755 | 3.12E-36 | 0.794 | 6.57E-85 | 0.695 | 1.33E-72 | 0.323 | 2.93E-15 |
| | **BD** | | **NEU** | | **CP** | | **ADHD** | |
| **F44** | 0.398 | 3.3E-10 | 0.414 | 9.80E-13 | 0.745 | 2.38E-11 | 0.608 | 5.05E-18 |
| **F45** | 0.32 | 5.99E-11 | 0.523 | 2.33E-45 | 0.57 | 1.75E-15 | 0.446 | 2.72E-22 |
| | **RA** | | **SLE** | | **SS** | | **SjS** | |
| **F44** | 0.193 | 0.0012 | 0.176 | 6.09E-02 | 0.365 | 0.0051 | 0.3 | 0.044 |
| **F45** | 0.104 | 0.0414 | 0.195 | 0.00420 | 0.283 | 1.00E-03 | 0.374 | 0.0006 |
| | **UC** | | **CD** | | **MS** | | **PS** | |
| **F44** | -0.038 | 0.61 | 0.1326 | 0.0251 | 0.245 | 0.0017 | 0.189 | 0.0136 |
| **F45** | 0.116 | 0.035 | 0.11 | 2.40E-02 | 0.118 | 0.0123 | 0.123 | 0.017 |
| | **GGE** | | **FE** | | **MIG** | | **IBS** | |
| **F44** | 0.097 | 0.178 | 0.277 | 0.0596 | 0.39 | 5.48E-9 | 0.605 | 3.32E-14 |
| **F45** | 0.038 | 0.514 | 0.21 | 0.0664 | 0.41 | 9.35E-19 | 0.648 | 8.30E-34 |
| | **T1D** | | **AITD** | | | | | |
| **F44** | 0.091 | 0.173 | 0.175 | 1.60E-03 | | | | |
| **F45** | 0.058 | 0.186 | 0.186 | 2.3E-06 | | | | |

**Supplementary Table 4. The validation analysis in Sweden Schizophrenia (SCZ) Study. With bold font we showed different direction of effect.**

| **Lead SNP** | **A1/A2** | ***p*-value, discovery dataset** | **A1 effect direction, discovery dataset** | ***p*-value, SCZ Study** | **A1 effect direction, SCZ study** |
|---|---|---|---|---|---|
| rs7830497 | T/G | 3.9e-09 | negative | 0.83 | negative (beta = -0.22) |
| rs7817744 | T/C | 1.1e-08 | **negative** | 0.18 | **positive** (beta =0.18, SE=0.13) |
| rs2908882 | T/C | 1.1e-08 | positive | 0.76 | positive (beta=0.04, SE = 0.13) |
| rs7004685 | A/G | 1.9e-10 | **negative** | 0.63 | **positive** (beta=0.04, SE=0.08) |

**DBDS research consortium**

Jakob Bay, Department of Clinical Immunology, Zealand University Hospital, Køge, Denmark

Andrea Barghetti, Department of Clinical Immunology, Copenhagen University Hospital, Rigshospitalet, Copenhagen, Denmark

Mette Skou Bendtsen, Department of Clinical Immunology, Copenhagen University Hospital, Rigshospitalet, Copenhagen, Denmark

Jens Kjærgaard Boldsen, Department of Clinical Immunology, Aarhus University Hospital, Aarhus, Denmark

Søren Brunak , Novo Nordisk Foundation, Center for Protein Research, Faculty of Health and Medical Sciences, University of Copenhagen, Copenhagen, Denmark

Nanna Brøns, Department of Clinical Immunology, Copenhagen University Hospital, Rigshospitalet, Copenhagen, Denmark

Alfonso Buil Demur, Institute of Biological Psychiatry, Mental Health Centre, Sct. Hans, Copenhagen University Hospital, Roskilde, Denmark

Johan Skov Bundgaard, Department of Clinical Immunology, Copenhagen University Hospital, Rigshospitalet, Copenhagen, Denmark

Lea Arregui Nordahl Christoffersen, Department of Clinical Immunology, Zealand University Hospital, Køge, Denmark

Maria Didriksen, Department of Clinical Immunology, Copenhagen University Hospital, Rigshospitalet, Copenhagen, Denmark

Khoa Manh Dinh, Department of Clinical Immunology, Aarhus University Hospital, Aarhus, Denmark

Joseph Dowsett, Department of Clinical Immunology, Copenhagen University Hospital, Rigshospitalet, Copenhagen, Denmark

Christian Erikstrup, Department of Clinical Immunology, Aarhus University Hospital, Aarhus, Denmark

Josephine Gladov, Department of Clinical Immunology, Aarhus University Hospital, Aarhus, Denmark

Daniel Gudbjartsson, deCODE Genetics, Reykjavik, Iceland

Thomas Folkmann Hansen, Danish Headache Center, Department of Neurology, Copenhagen University Hospital, Rigshospitalet-Glostrup, Copenhagen, Denmark

Dorte Helenius Mikkelsen, Institute of Biological Psychiatry, Mental Health Centre, Sct. Hans, Copenhagen University Hospital, Roskilde, Denmark

Lotte Hindhede, Department of Clinical Immunology, Aarhus University Hospital, Aarhus, Denmark

Henrik Hjalgrim, Danish Cancer Society Research Center, Copenhagen, Denmark

Jakob Hjorth von Stemann, Department of Clinical Immunology, Copenhagen University Hospital, Rigshospitalet, Copenhagen, Denmark

Bitten Aagaard, Department of Clinical Immunology, Aalborg University Hospital, Aalborg, Denmark

Kathrine Kaspersen, Department of Clinical Immunology, Aarhus University Hospital, Aarhus, Denmark

Bertram Dalskov Kjerulff, Department of Clinical Immunology, Aarhus University Hospital, Aarhus, Denmark

Lisette Kogelman, Danish Headache Center, Department of Neurology, Copenhagen University Hospital, Rigshospitalet-Glostrup, Copenhagen, Denmark

Mette Kongstad, Department of Clinical Immunology, Copenhagen University Hospital, Rigshospitalet, Copenhagen, Denmark

Susan Mikkelsen, Department of Clinical Immunology, Aarhus University Hospital, Aarhus, Denmark

Christina Mikkelsen, Department of Clinical Immunology, Copenhagen University Hospital, Rigshospitalet, Copenhagen, Denmark

Line Hjorth Sjernholm Nielsen, Department of Clinical Immunology, Aarhus University Hospital, Aarhus, Denmark

Janna Nissen, Department of Clinical Immunology, Copenhagen University Hospital, Rigshospitalet, Copenhagen, Denmark

Mette Nyegaard, Department of Health Science and Technology, Faculty of Medicine, Aalborg Univeristy, Aalborg, Denmark

Sisse Rye Ostrowski, Department of Clinical Immunology, Copenhagen University Hospital, Rigshospitalet, Copenhagen, Denmark

Frederikke Byron Pedersen, Department of Clinical Immunology, Copenhagen University Hospital, Rigshospitalet, Copenhagen, Denmark

Ole Birger Pedersen, Department of Clinical Immunology, Zealand University Hospital, Køge, Denmark

Liam James Elgaard Quinn, Department of Clinical Immunology, Zealand University Hospital, Køge, Denmark

Þórunn Rafnar, deCODE Genetics, Reykjavik, Iceland

Palle Duun Rohde, Department of Health Science and Technology, Faculty of Medicine, Aalborg Univeristy, Aalborg, Denmark

Klaus Rostgaard, Danish Cancer Society Research Center, Copenhagen, Denmark

Andrew Joseph Schork, Institute of Biological Psychiatry, Mental Health Centre, Sct. Hans, Copenhagen University Hospital, Roskilde, Denmark

Michael Schwinn, Department of Clinical Immunology, Copenhagen University Hospital, Rigshospitalet, Copenhagen, Denmark

Erik Sørensen, Department of Clinical Immunology, Copenhagen University Hospital, Rigshospitalet, Copenhagen, Denmark

Kari Stefansson, deCODE Genetics, Reykjavik, Iceland

Hreinn Stefánsson, deCODE Genetics, Reykjavik, Iceland

Jacob Træholt, Department of Clinical Immunology, Copenhagen University Hospital, Rigshospitalet, Copenhagen, Denmark

Unnur Þorsteinsdóttir , deCODE Genetics, Reykjavik, Iceland

Mie Topholm Bruun, Department of Clinical Immunology, Odense University Hospital, Odense, Denmark

Henrik Ullum, Statens Serum Institut, Copenhagen, Denmark

Thomas Werge, Institute of Biological Psychiatry, Mental Health Centre, Sct. Hans, Copenhagen University Hospital, Roskilde, Denmark
